# Language-Related Disparities in History Documentation in Patients Admitted for Heart Failure

**DOI:** 10.64898/2026.05.19.26353593

**Authors:** Eric Raphael Gottlieb, Irene Dankwa Mullan, Leo Anthony Celi

**Affiliations:** Department of Medicine, Cambridge Health Alliance, Cambridge, MA; Harvard Medical School, Boston, MA; Laboratory for Computational Physiology, Massachusetts Institute of Technology, Cambridge, MA; Milken Institute School of Public Health, George Washington University, Washington DC; Division of Pulmonary, Critical Care and Sleep Medicine, Beth Israel Deaconess Medical Center, Boston, MA; Department of Biostatistics, Harvard T.H. Chan School of Public Health, Boston, MA

**Keywords:** Translation, Interpreter Services, Documentation, Health Equity

## Abstract

**Introduction:** Patients hospitalized with heart failure who do not speak English as their primary language face communication barriers, however the impact on documented History of Present Illness (HPI) and Review of Systems (ROS) has not been reported.

**Methods:** This retrospective cohort study was based on MIMIC-IV, an anonymized clinical database. Adult patients admitted to general medicine or cardiology services with heart failure (by DRG) were identified. Multivariable linear regression was used to assess for an association between language (English vs. non-English) and word counts for HPI+ROS and HPI word counts. Qualitative differences in texts were also analyzed using Claude Opus 4.6.

**Results:** In a cohort of 552 patients, non-English language (N = 81) was associated with a shorter HPI+ROS (coef. -33.387, 95% CI [-62.076, -4.697], p = 0.023) controlling for age (coef. -1.023, 95% CI [-1.817, -0.230], p = 0.012) and Elixhauser score (coef. 10.391, 95% CI [7.078, 13.705], p<0.001). Similar associations were found for HPI alone. Qualitative differences included less discussion of symptoms and timing of onset.

**Discussion:** HPI+ROS and HPI were more abbreviated when the primary documented language was not English. This has important implications for equitable care and the development of emerging translation and documentation technologies.

## Background

Heart failure is a clinical diagnosis made on the basis of characteristic signs and symptoms, such that obtaining and recording a complete and accurate narrative History of Present Illness (HPI) and structured Review of Systems (ROS) at the time of hospital admission are of paramount importance. For patients who face language barriers, this encounter is qualitatively different, and despite governmental and accrediting bodies requiring hospitals to provide interpreter services,^1^ multiple constraints remain, including but not limited to time, quality of and access to interpreters, and history obtained indirectly through third parties.^2,3^

Although impacts of language barriers^4^ and a wide variety of quality improvement interventions have been studied,^5^ the objective impact on formal documented HPI has not been reported. Meanwhile, ambient artificial intelligence is transforming this initial encounter,^6^ potentially leaving these patients even further behind. This emphasizes the need for a baseline understanding of how language barriers impact documented histories. In this study, we assessed documentation disparities in length and quality of the HPI and ROS written for patients admitted for heart failure depending on whether or not English was their documented primary language.

## Methods

We conducted a retrospective observational cohort study using the Medical Information Mart for Intensive Care (MIMIC-IV, Version 3.1).^7,8^ MIMIC was approved for research by the institutional review boards of Beth Israel Deaconess Medical Center (BIDMC) in Boston, MA (2001-P-001699/14) and the Massachusetts Institute of Technology (0403000206) without a requirement for individual patient informed consent because it is deidentified and publicly available. This study followed the STROBE guidelines for observational studies.^9^

Data were extracted and analyzed using the Google BigQuery (Alphabet Inc.) cloud platform and RStudio Version 2025.09.2 (Posit Software) with the R 4.5.2 programming language (R Foundation for Statistical Computing), respectively.

We analyzed inpatient hospitalization discharge summaries, which typically include the initial HPI and ROS, and matched them to records of hospital admissions. Only adult patients with a diagnosis related group (DRG) matching “HEART FAILURE” who were admitted to either the cardiology or general medicine services were included in this study. To obtain what is likely the most complete initial history, for each patient the chronological first heart failure admission in the data set was selected. HPI and ROS were extracted from the discharge summaries by identifying a block of text beginning with the words “History of Present Illness” and ending with “ROS” to include only the HPI, or ending with “Past Medical History” to obtain a combination of the HPI and ROS. Primary language of English versus Other Language was determined from a structured field in the “Admissions” table.

Demographic and clinical variables from the time of admission were extracted. International Classification of Diseases (ICD) codes were used to calculate unweighted Elixhauser comorbidity scores^10^ using the R ‘comorbidity’^11^ package. Multivariable linear regression models were constructed to assess for the association between non-English language and word count of HPI+ROS and HPI, respectively. Patient age and Elixhauser score were found in initial analyses to be significant covariates and were included in the final models, while other demographic variables were explored and found to be nonsignificant, including race, gender, insurance, and marital status. Interactions between race and English language were ruled out as well to avoid confounding between language and race/ethnicity. A p-value of <0.05 was considered statistically significant. For visual presentation, a subset of equal numbers of patients matched according to the significant variables was extracted using the R ‘MatchIt’ package.^12^

To complement length-based measures with an assessment of content quality and narrative complexity, we conducted a large language model (LLM)-assisted comparison of HPI and ROS stratified by language. HPIs were uploaded to the Claude Opus 4.6 AI model (Anthropic PBC) using a prespecified prompt to identify qualitative differences between the text as written for English and non-English-speaking patients. The use of an external model complied with MIMIC data use policy requirements ensuring that the AI system did not retain submitted data for future model training.

## Results

There were 552 patients included in this study, of whom 81 had primary languages other than English, the most common being Russian, Spanish, and Chinese. These patients were older, less likely to be single, more likely to be on Medicaid, and less likely to have private insurance. More than 90% of patients in both groups were admitted from the Emergency Department and similar proportions were admitted at night (between 7:00pm and 6:59 am). Demographic and clinical characteristics, word counts, as well as in-hospital death and length of stay are shown in Table 1. A distribution of word counts for a matched subset of patients is shown in Figure 1.

**Table 1:** Characteristics of Subjects and Outcomes.

| Variable | Overall | English | Non-English | Missing (%) |
| --- | --- | --- | --- | --- |
| N | 552 | 471 | 81 |  |
| Age (Years) (Mean/SD) | 76.2 (12.8) | 75.5 (13.1) | 80.3 (10.2) | 0.0 |
| Male (%) = M | 237 (42.9) | 204 (43.3) | 33 (40.7) | 0.0 |
| Race (%) |  |  |  | 0.0 |
| Asian | 21 (3.8) | 7 (1.5) | 14 (17.3) |  |
| Black | 97 (17.6) | 89 (18.9) | 8 (9.9) |  |
| Hispanic | 13 (2.4) | 2 (0.4) | 11 (13.6) |  |
| Other/Unknown | 28 (5.1) | 19 (4.0) | 9 (11.1) |  |
| White | 393 (71.2) | 354 (75.2) | 39 (48.1) |  |
| Marital Status (%) |  |  |  | 0.5 |
| Divorced | 48 (8.7) | 43 (9.2) | 5 (6.2) |  |
| Married | 220 (40.1) | 183 (39.0) | 37 (46.2) |  |
| Single | 120 (21.9) | 112 (23.9) | 8 (10.0) |  |
| Widowed | 161 (29.3) | 131 (27.9) | 30 (37.5) |  |
| Language (%) |  |  |  | 0.0 |
| Chinese | 13 (2.4) | 0 (0.0) | 13 (16.0) |  |
| English | 471 (85.3) | 471 (100.0) | 0 (0.0) |  |
| Other Language | 22 (4.0) | 0 (0.0) | 22 (27.2) |  |
| Russian | 33 (6.0) | 0 (0.0) | 33 (40.7) |  |
| Spanish | 13 (2.4) | 0 (0.0) | 13 (16.0) |  |
| Insurance (%) |  |  |  | 0.9 |
| Medicaid | 49 (9.0) | 33 (7.1) | 16 (20.0) |  |
| Medicare | 433 (79.2) | 372 (79.7) | 61 (76.2) |  |
| Other Insurance | 3 (0.5) | 3 (0.6) | 0 (0.0) |  |
| Private | 62 (11.3) | 59 (12.6) | 3 (3.8) |  |
| Night Admission (%) = 1 | 276 (50.0) | 237 (50.3) | 39 (48.1) | 0.0 |
| ED Admit (%) = 1 | 502 (90.9) | 426 (90.4) | 76 (93.8) | 0.0 |
| Elixhauser Score (Mean/SD) | 2.0 (3.0) | 2.0 (3.0) | 2.4 (3.1) | 0.0 |
| Sodium (mEq/L) [Median/IQR] | 140.0 [136.8, 142.0] | 140.0 [137.0, 142.0] | 138.0 [136.0, 143.0] | 4.3 |
| Potassium (mEq/L) [Median/IQR] | 4.2 [3.8, 4.5] | 4.2 [3.8, 4.5] | 4.1 [3.8, 4.5] | 3.8 |
| Bicarbonate (mEq/L) [Median/IQR] | 27.0 [23.0, 30.0] | 27.0 [23.0, 30.0] | 26.0 [24.0, 30.0] | 4.3 |
| BUN (mg/dL) [Median/IQR] | 27.0 [19.0, 43.0] | 28.0 [19.0, 43.0] | 26.0 [18.0, 47.0] | 4.5 |
| Creatinine (mg/dL) [Median/IQR] | 1.3 [1.0, 1.9] | 1.3 [1.0, 1.9] | 1.2 [1.0, 1.9] | 4.3 |
| Hemoglobin (g/dL) [Median/IQR] | 10.4 [9.0, 11.9] | 10.4 [9.0, 11.8] | 10.6 [9.1, 12.2] | 6.9 |
| HPI+ROS Length (Words) [Median/IQR] | 293.5 [230.5, 376.0] | 298.0 [231.0, 385.0] | 285.0 [226.0, 334.0] | 0.0 |
| HPI Length (Words) [Median/IQR] | 260.0 [198.0, 335.0] | 262.0 [198.5, 340.0] | 247.0 [197.0, 300.0] | 0.0 |
| ROS Word Count (Words) [Median/IQR] | 26.0 [11.8, 48.2] | 26.0 [12.0, 49.5] | 25.0 [8.0, 40.0] | 0.0 |
| Length of Stay (Days) (Mean/SD) | 5.6 (3.9) | 5.6 (3.9) | 5.5 (4.2) | 0.0 |
| Died (%) = 1 | 14 (2.5) | 9 (1.9) | 5 (6.2) | 0.0 |

**Figure 1.**
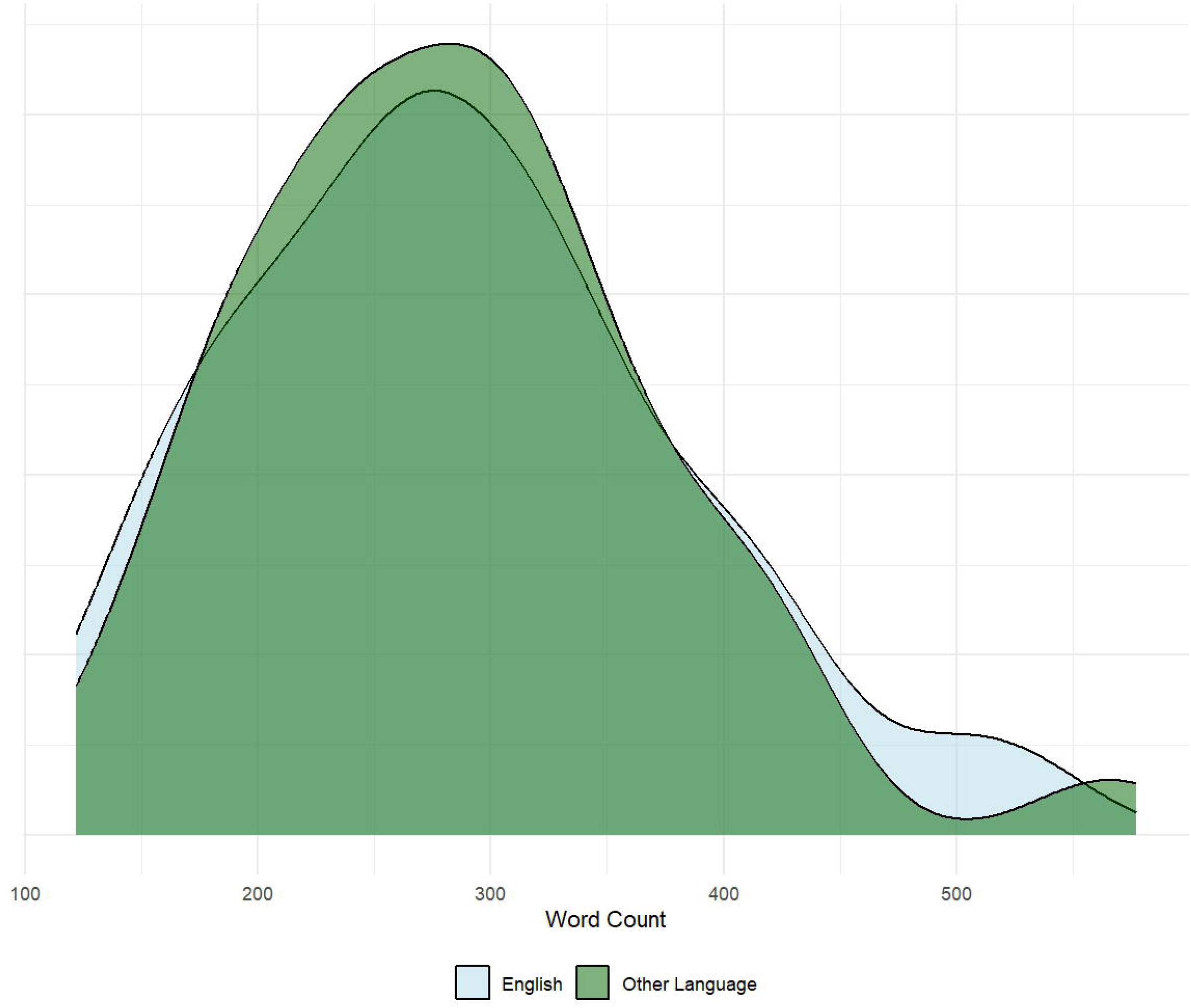
Title: Distribution of Word Counts for a Subset of Matched Patients with English and Non-English Primary Language.

In a multivariable linear regression, having a primary language other than English was associated with a shorter HPI+ROS word count (coef. -33.387, 95% CI [-62.076, - 4.697], p = 0.023) when controlling for age by year (coef. -1.023, 95% CI [-1.817, - 0.230], p = 0.012) and Elixhauser score (coef. 10.391, 95% CI [7.078, 13.705], p<0.001). Similar associations were found for HPI alone with language other than English (coef. -29.653, 95% CI [-58.057, -1.250], p = 0.041), again controlling for age (coef. -1.033, 95% CI [-1.819, -0.248], p = 0.010) and Elixhauser score (coef. 11.226, 95% CI [7.946, 14.507], p<0.001).

AI analysis of the text identified a number of characteristic differences between documentation in non-English and English classified patients, including less discussion of ancillary symptoms such as fevers and urinary and other respiratory symptoms and symptom timing. The entire readout is presented in Supplement 1.

## Discussion

In this study, we showed that when controlling for patient age and comorbidity burden, a documented language other than English was associated with shorter, less comprehensive and less symptom-focused written histories in patients admitted for heart failure.

While a longer history is not inherently superior and our analysis of the notes is limited, our findings are some of the first objective evidence of an inconsistency in history-taking, which is considered by cardiology society guidelines to be a patient safety risk.^13^ Furthermore, given that patients classified as non-English-speaking were older, had more comorbidities, and likely faced additional adverse social hurdles, this brevity is unlikely to be justified by a simpler clinical presentation and suggests potentially problematic disparities in how the history is gathered and recorded. For example, the sparser discussion of fevers and urinary symptoms could lead to delayed detection of urinary sepsis in a patient admitted for heart failure, and the lack of interrogation of other respiratory symptoms may limit consideration of competing diagnoses besides heart failure. As a result, language barriers may lead to incorrect or missed diagnoses.

A strength of this study is that because it was limited to a single primary admission diagnosis of heart failure, confounding factors were minimized. We also made conservative use of AI to better understand the qualitative differences beyond simple word count.

Limitations include first that language information is limited to one structured field, which we further stratified to English versus non-English, and may or may not be the language used during the interaction. Second, this study is limited to a single health system with patient and staff characteristics and practices that may not be externally generalizable. Third, although we adjusted for available patient and admission characteristics, residual confounding is likely. Fourth, HPI and ROS were taken from discharge summaries and could have included documentation elements not strictly related to narrative history or that were revised from the initial H&P. Finally, large language model (LLM) analysis is inherently affected by the choice of model and prompting, and its findings were not subjected to formal statistical analysis.

## Conclusion

In a DRG-defined heart failure cohort, formal documented histories were more abbreviated when the primary documented language was not English. Further research is needed to validate these findings in other settings and diagnoses, further characterize the disparities in documented histories, and identify ways to mitigate them.

## Data Availability

MIMIC is publicly available with training in human subjects research and application. Statistical code is available upon request.

https://physionet.org/content/mimiciv/3.1/

