## Supplement 1 for "Language-Related Disparities in History Documentation in Patients Admitted for Heart Failure"

**Claude AI Readout Comparing English to non-English-Speaking Patients**

### Sample sizes

| **Group** | **N** | **% of Total** |
| --- | --- | --- |
| English-Primary | 471 | 85.3% |
| Other Language | 81 | 14.7% |
| **Total** | **552** | **100%** |

471 English-Primary vs. 81 Other Language patients.

### Documentation & Communication

The most striking difference is language documentation: 27% of Other Language notes explicitly reference the patient “speaking” a non-English language, and 6% note an interpreter was used — compared to essentially 0% in the English group. Notes for Other Language patients are also somewhat shorter on average (281 vs. 315 words), suggesting that language barriers may lead to briefer or less detailed documentation.

### History of Present Illness (HPI)

#### More prominent in other_language=1:

- Hypertension is mentioned significantly more often (62% vs. 50%), suggesting HTN is a more prominent part of the presenting picture or past medical history.
- References to pulmonary findings, heart failure, and cardiac terms (pulmonary, effusion, heart, diastolic, ischemic, troponin) are modestly more frequent.
- Intermittent symptoms and bilateral findings are noted more often.
- Moderate severity descriptors appear more.

#### More prominent in other_language=0:

- Temporal and symptomatic detail is richer: words like increased, during, daily, weeks, time, currently, prior, acute appear more often — indicating more granular history-taking with clear timelines.
- COPD is much more frequently documented (16% vs. 6%), perhaps reflecting a real prevalence difference or differential access to prior diagnosis.
- Urinary symptoms are notably more prominent (54% vs. 42%), including dysuria, frequency, and urinary as discrete terms — possibly reflecting more thorough genitourinary ROS.
- Exertional symptoms (DOE, 44% vs. 36%) and orthopnea (48% vs. 41%) appear more frequently.
- Sputum production and wheezing are more often documented.
- Fevers/chills are documented more (79% vs. 72%).

### Review of Systems (ROS)

- The use of the word “denies” (a standard ROS negation phrase) is slightly lower in Other Language notes (74% vs. 80%), which may reflect that a structured, comprehensive ROS is harder to document when a language barrier exists.
- Core symptoms like shortness of breath, chest pain, nausea, and dizziness are documented at nearly identical rates, suggesting these cardinal symptoms are captured regardless of language.
- Urinary ROS is notably less complete in Other Language notes (-12%).
- Fever/chills and exertional symptoms are less commonly documented in Other Language notes.

### Summary Interpretation

The differences suggest that language barriers are associated with less detailed and granular documentation — particularly for nuanced symptoms (exertional, urinary, respiratory quality), temporal patterns, and structured ROS negations. The core chief complaint is captured similarly in both groups, but the surrounding clinical context and review of systems is somewhat thinner in Other Language notes. This is consistent with known challenges in history-taking across language barriers, even with interpreter use.
